# An algorithm for automated detection of evoked potentials from polarity reversed electrical stimulations

**DOI:** 10.1101/2023.05.09.23289729

**Authors:** Jessica S L Vidmark, Terence D Sanger

## Abstract

**Background:** In neurophysiological research involving neural responses to electrical stimuli, each recording must be searched for evoked potentials (EPs) prior to further analysis. Conducting this process manually is time consuming for the researcher and also inserts bias. However, automated detection methods often struggle to distinguish between artifacts and neural responses, which can have highly complex and varying shapes.

**Methods:** We have developed a novel algorithm for automated detection of polarity reversed EPs (ADPREP), which uses the knowledge that reversing the polarity of a pair of stimulating contacts reverses the sign of the stimulus artifact, but not that of the neural response. Hence, our algorithm searches for any positive correlation between the recordings from two polarity reversed stimulations, after removing any stimulus decay artifacts. If the peak-to-peak amplitude in a positively correlated region surpasses a user-defined threshold, the recording is labeled as an EP; otherwise, it is not. Neural recordings from deep brain nuclei and cortical regions during deep brain stimulation (DBS) in 28 pediatric patients with dystonia were used to test and prove the validity of the method.

**Results:** The ADPREP algorithm is able to distinguish EPs of varying shapes and sizes with a high level of accuracy, as early as 0.35 ms after stimulation, despite large stimulus artifacts. The algorithm has proven useful in initial tests with DBS data in hundreds of stimulation/recording combinations within the basal ganglia and thalamic nuclei as well as from these deep brain nuclei to cortex, at stimulation frequencies up to 250 Hz.

**Conclusion:** Our automated EP detection algorithm can accurately detect DBS EPs in deep brain nuclei and cortex, and has promising applications in other stimulation and recording modalities that allow for polarity reversal of the recorded stimulus artifact. The algorithm successfully labels EPs of varying shapes and sizes, as early as a fraction of a millisecond after stimulation, in a range of stimulus frequencies and stimulation-recording pairs – even under large stimulus decay artifacts and in same-lead stimulation and recordings. As such, it is a great method to improve efficacy and minimize human bias, setting up for more reliable conclusions to be drawn from the data.

## Introduction

Deep brain stimulation (DBS) is a common treatment of movement disorders [1]–[5]. When used in combination with externalized depth electrodes, which can both stimulate and record, valuable insight can be obtained into the brain’s neural responses to electrical stimulation (referred to as evoked potentials or EPs) [6]–[8]. With the numerous combinations of possible stimulation and recording locations, stimulation pattern and frequency, and other parameters, thousands of connections can be studied—all of which must be searched for EPs prior to further analysis and conclusions. Conducting this process manually is time consuming for the researcher, and also inserts bias. However, automated detection methods often struggle to distinguish between artifacts and neural responses, which can have highly complex and varying shapes.

One experimental approach to distinguishing stimulus artifacts from neural responses is the use of polarity-reversed stimulations, wherein electrical stimulation is performed through a pair of contacts (a & b) in both polarities: with a as the cathode and b as the anode (“cathodic stimulation”), and vice versa (“anodic stimulation”) [8], [9]. The method is useful because flipping the cathodic and anodic stimulations contacts reverses the sign of the stimulus artifact, but not the neural response [9]; hence, neural responses are expected to positively correlate between the cathodic and anodic recordings, while artifactual components would correlate negatively, or not at all.

I have developed a novel algorithm for automated detection of polarity reversed EPs (ADPREP). This method uses the knowledge that in a pair of recordings during polarity reversed (cathodic and anodic) stimulations, neural responses would show up as positive correlations between the cathodic and anodic recordings, after removal of stimulus artifacts. Hence, the ADPREP method starts by detecting and removing decay artifacts after the main stimulus artifact by fitting to a sum of two exponential decays for each of the two stimulation types. If a significantly positive correlation is found between the resulting artifact-reduced cathodic and anodic recordings, and the amplitude of the resulting average response in this correlated region is greater than a user-defined threshold, the recording is labeled as an EP; otherwise, it is not. The EPs can thereafter be characterized by features such as their peak-to-peak amplitude (P2P), time-to-(first-)peak delay (T2P), frequency components, and more.

Initial results are promising, proving the ADPREP algorithm’s ability to distinguish EPs of many varying shapes and sizes from stimulus artifacts and other noise in the recordings, saving time for the user and minimizing human bias. One of the benefits of this method is that it does not require the shape of the EP to be pre-defined; because of this, the algorithm does not discriminate towards or against any EP shapes. Another benefit is its user-defined amplitude threshold, which can be lowered or raised to reduce the false negative or false positive rates, respectively, to match the preference of the researcher or project.

## Materials and Methods

The main inputs to the ADPREP method are recordings obtained during polarity reversed stimulations, from electrodes situated close enough to the stimulating electrode pair to experience a reversal of the stimulus artifact between the two stimulation types (cathodic and anodic). The algorithm, which is outlined in Figure 1, starts by removing any decay artifacts from the recordings of each stimulus type, after the stimulus artifact. Next, the correlation between the resulting artifact-reduced cathodic and anodic recordings is evaluated using moving correlation windows. If a significantly positive correlation is detected above a user-defined threshold, the average response between the cathodic & anodic recordings is obtained, and the P2P amplitude within this positive correlation region is determined. Finally, if this P2P amplitude surpasses an empirically chosen threshold, the average recording is labeled as an EP; otherwise, it is not.

**Figure 1.**
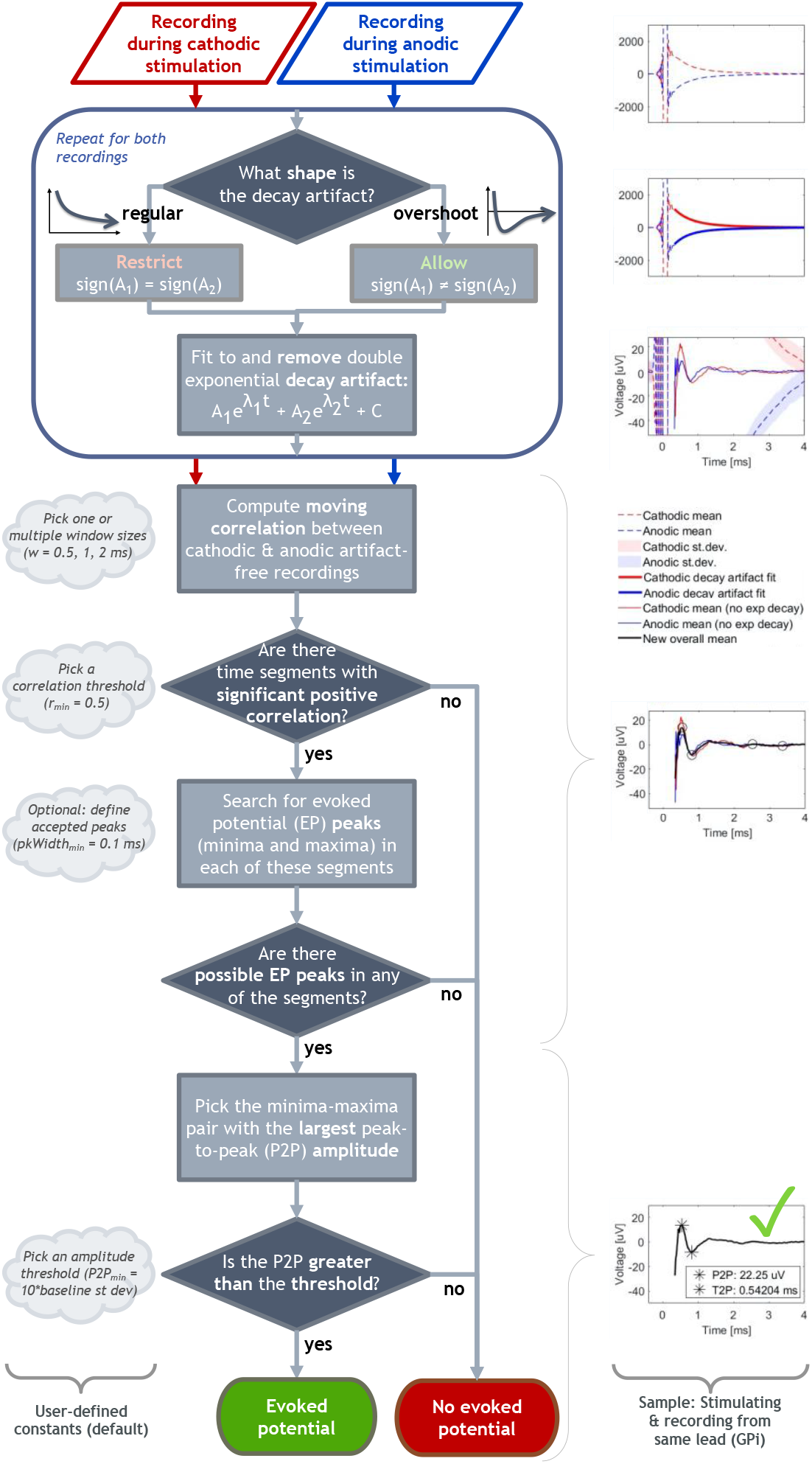
Flowchart of the ADPREP algorithm (left) with examples visualizing the main steps (right).

For a full list of the parameter values, initial conditions, and other restrictions used in our dataset, see Table 1 & Table 2.

**Table 1.** ADPREP parameter values used for the dataset presented in this paper.

| Parameter | Description | Unit | Our value(s) |
| --- | --- | --- | --- |
| w | Moving correlation window size(s) | ms | [0.5, 1, 2] |
| $r_{\min}$ | Correlation threshold (minimum) | - | 0.5 |
| $A_{\min}$ | Amplitude threshold (minimum) | uV | 10x baselineStd* |

\*We defined our "baseline standard deviation" (baselineStd) as the standard deviation of the detrended average signal of the cathodic & anodic stimulus-triggered averages from 1.4 to 0.3 ms before the stimulus artifact.

**Table 2.** SEDAM parameter initial values, lower bounds, and upper bounds used for the dataset presented in this paper. Values in italicized brackets denote the values when the decay artifact fit is forced to be a “simple” exponential, by making the amplitudes of the exponential decays have the same sign (note that sign(A_1_) is multiplied by A_2_ in this restricted case to force this relationship). Otherwise, the values chosen are the same for both the restricted (forced “simple” exponential decay model) and unrestricted cases (both “simple” and “overshoot” exponential decays allowed).

| Parameter | Description | Unit | Initial value | Lower bound | Upper bound |
| --- | --- | --- | --- | --- | --- |
| $A_1$ | Amplitude of exponential decay 1 | uV | $\text{AMDS} \cdot \text{sign}(\text{dataSeg}(\text{ind}_{\text{ADMS}}))$ | -4*AMDS<br>[-2*AMDS] | 4*AMDS<br>[2*AMDS] |
| $A_2$ | Amplitude of exponential decay 2 | uV | $-\text{AMDS} \cdot \text{sign}(\text{dataSeg}(\text{ind}_{\text{ADMS}}))$<br>[AMDS] | -4*AMDS<br>[0] | 4*AMDS<br>[2*AMDS] |
| $\lambda_1$ | Time constant of exponential decay 1 | 1/ms | -5 | -12 | -0.01 |
| $\lambda_2$ | Time constant of exponential decay 2 | 1/ms | -1 | -12 | -0.01 |
| C | Baseline offset | uV | $\text{dataSeg}(\text{end})$ | $-\text{abs}(\text{dataSeg}(\text{end}))$ | $\text{abs}(\text{dataSeg}(\text{end}))$ |

Abbreviations & variables: abs, absolute value; AMDS, absolute max of the data segment (dataSeg); dataSeg, data segment used to fit (and remove) decay artifact and subsequently search for an EP; end, last index in segment; $\text{ind}_{\text{ADMS}}$ , index where AMDS occurs.

### Valid data

To ensure the validity of the assumptions of the polarity reversal approach used in this algorithm, the neurophysiological recordings must be obtained from electrodes situated close enough to the stimulating electrode pair to experience a reversal of the stimulus artifact between the two stimulation types (cathodic and anodic). Examples of such valid recording & stimulation combinations include recording from deep brain nuclei (e.g., using sEEG) or cortical regions (e.g., using electroencephalography, EEG) during simultaneous deep brain stimulation. Examples from the former are presented in this paper.

Note that if the recorded data’s signal-to-noise ratio is acceptable, single trial recordings may be used. Otherwise, stimulus-triggered averaging can be implemented to increase SNR prior to using the ADPREP algorithm.

### Decay artifact removal

The polarity reversal approach builds on the assumption that when the polarity of the stimulating contact pair is switched, the polarity of the stimulus artifact is reversed, whereas that of the neural response is not. While the main stimulus artifact can be relatively brief and return to baseline prior to the start of the evoked response, so-called decay artifacts may extend for a longer period, distorting the neural response [10]–[12]. Just like the main stimulus artifact, the polarity of the decay artifact depends on the polarity of the stimulation. Because of these opposing polarities of the decay artifacts from the polarity reversed stimulations, they can make the correlation between the cathodic and anodic recordings seem non-significant or negative, even if there is an underlying neural response that is common to both recordings. Therefore, these decay artifacts must be removed prior to the ADPREP algorithm’s correlation analysis.

The decay artifact removal is based on an approach that fits the data to the sum of two double exponentials [12]:

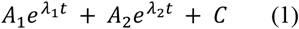

where *A*_*1*_ and *A*_*2*_ are the amplitudes and *λ*_*1*_ and *λ*_*2*_ the decay constants of the two exponential components, and *C* is the baseline offset.

In our data, we have encountered not only “regular” exponential decay artifacts, which decay towards 0 without crossing the x-axis, but also decay artifacts that “overshoot” the x-axis before stabilizing back towards 0 (Figure 1, top). The SEDAM model (Equation 1) can model both these types of decay artifacts: when *sign(A*_*1*_) = *sign(A*_*2*_), the resulting SEDAM decay artifact is “regular”, whereas when *sign(A*_*1*_) ≠ *sign(A*_*2*_), an “overshoot” decay artifact is modelled.

However, due to the regression’s high sensitivity to the initial values of *A*_*1*_ and *A*_*2*_, two separate iterations of the regression are run. First, the initial values of *A*_*1*_ and *A*_*2*_ have the same sign, and the upper and lower boundaries of the coefficients are chosen to force *sign(A*_*1*_) = *sign(A*_*2*_). In the second iteration, the initial values of *A*_*1*_ and *A*_*2*_ are of opposite signs, and the relation between the signs of *A*_*1*_ and *A*_*2*_ is not restricted (i.e., *sign(A*_*1*_) ≠ *sign(A*_*2*_) and *sign(A*_*1*_) = *sign(A*_*2*_) are both allowed). The parameters from the regression that generated the best fit (defined by the R^2^ value) are used to model the decay artifact and subtract it from the recording.

This process is repeated for both the cathodic and anodic signals, to generate artifact-reduced recordings for use in the subsequent steps of the ADPREP algorithm.

### Moving correlation

After decay artifact removal, the artifact-reduced post-stimulation recordings are used to determine the correlation between the cathodic and anodic signals. This main part of the ADPREP algorithm is built on the assumption that a neural response evoked by a stimulation of a pair of nearby contacts would be similarly elicited if the polarity of the stimulating contact pair was reversed (i.e., cathodic vs anodic stimulation). Hence, any neural response to the stimulation should display a significant positive correlation between the artifact-reduced polarity reversed recordings.

Since neural responses can be brief and may not span the whole recording, *moving* correlation is required (as opposed to determining the correlation between the signals over the entire recording). We use the MATLAB (The MathWorks, Inc., Natick, MA, USA) File Exchange function *movcorr* [13] to calculate the correlation between the artifact-reduced cathodic and anodic recordings over time; in a similar manner to moving average, it calculates the Pearson correlation in a user-defined window at each point as it moves over the x-axis, one sample at a time. This allows us to find local correlations that may be representative of neural responses evoked by the electrical stimulation.

As neural responses can have varying frequencies, the width of the moving correlation window should be adjusted accordingly. The ADPREP algorithm allows for multiple user-defined window sizes, which should be selected based on the expected frequency and duration of the neural response. The moving correlation is calculated over the recording after the stimulation, for each of the window sizes. The time regions that display significant positive correlation above a user-defined threshold of Pearson’s *r* – using any or all of the window sizes – will be searched for possible EPs.

### Peak detection and amplitude threshold

The possible EP regions located using the above methods are checked for EPs using the MATLAB *findpeaks* function. In the ADPREP algorithm, the function is simply used to find the positive and negative peaks with the largest and smallest values, respectively, in each potential EP region. Further *findpeaks* restrictions, such as *minPeakWidth* or *minPeakHeight*, are optional.

After finding the peaks in each possible EP region, the region with the greatest P2P amplitude is assumed to contain the (main) neural response. In the final step, the P2P amplitude is compared to a user-defined minimum amplitude threshold, which can be set to a firm value (e.g., 10 uV) or require a certain SNR (as in our sample dataset, Table 1). If the P2P surpasses the amplitude threshold, the located peaks are labeled as representative of an EP. However, if any of the above thresholds were not met, the recording is labeled as *not* containing an EP.

### Sample data

In our lab, we work with pediatric patients with dystonia to study, among other things, how neural responses evoked by thalamic and basal ganglia DBS spread throughout deep brain regions and to the cortex [6], [14]. As such, the data used in this paper to present sample results from the ADPREP algorithm are bipolar recordings from sEEG micro-electrodes (Adtech MM16C, Adtech Medical Instrument Corp., Oak Creek, WI, USA) in deep brain structures (thalamus and basal ganglia) during DBS (through the sEEG leads) of thalamic or basal ganglia nuclei, from 28 subjects. These subjects, or their legal guardians, provided HIPAA authorization for research use of protected health information prior to participation. In addition, written informed consent was given for surgical procedures conforming to standard hospital practice and for research use of electrophysiological data. This study conformed to the ethical guidelines of the Declaration of Helsinki, and the institutional review boards of Children’s Hospital Los Angeles (CHLA) and Children’s Health Orange County (CHOC) approved all research use of data.

## Results

The ADPREP algorithm is able to distinguish EPs of varying shapes and sizes with a high level of accuracy, as early as 0.35 ms after stimulation, despite large stimulus artifacts (even in same-lead stimulation & recordings). The algorithm has proven useful in its initial tests during DBS in hundreds of stimulation/recording combinations within the basal ganglia and thalamic nuclei (sEEG recordings) and from these deep brain nuclei to cortex (EEG recordings), during stimulation frequencies up to 250 Hz, in 28 subjects to date. Sample results are displayed in Figure 2, selected to show the wide range of EPs that the algorithm can detect (even under large decay artifacts) and exemplify rejected recordings that may have been labeled as EPs by methods that only use the cathodic and anodic signals independently, or the average between them.

**Figure 2.**
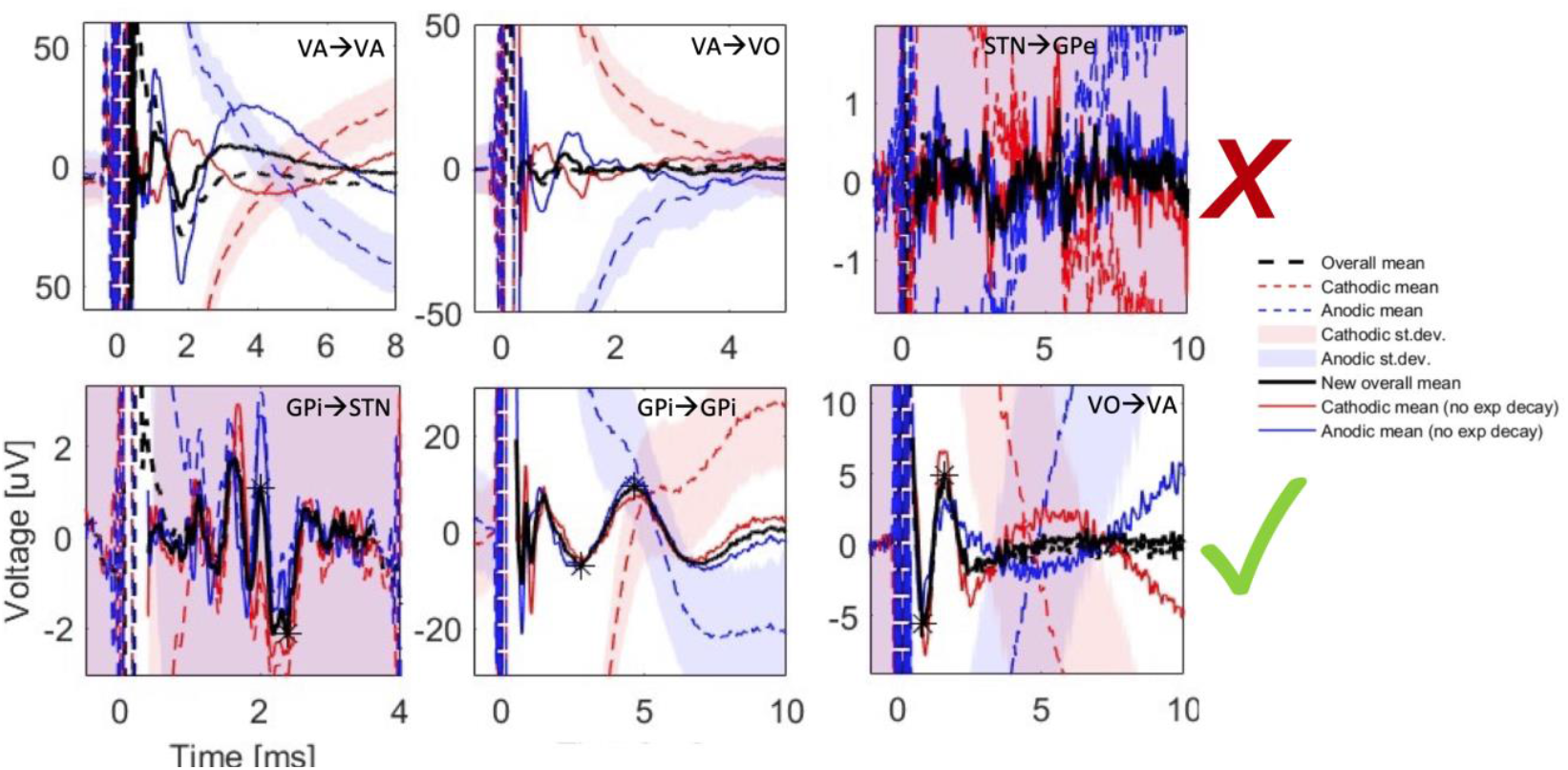
Examples of rejected and detected EPs using the ADPREP algorithm. The top and bottom rows show sample rejected and detected EPs, respectively, using the user-defined variables listed in Table 1 & Table 2. Dashed red and blue lines are stimulus-triggered averages of the cathodic and anodic stimulation types, respectively, with shaded standard deviations; dashed black lines are the average between the original cathodic and anodic signals. Similarly, solid red and blue lines are the signals after removal of decay artifacts; solid black lines are the average between the decay artifact-reduced cathodic and anodic signals. Stars denote the detected positive and negative peaks of the labeled evoked potentials. All examples are taken from different subjects and stimulation/recording combinations, as denoted in the annotation in each subplot (stimulation → recording region). Y-axis: recorded voltage [uV]; x-axis: time after stimulation [ms].

## Discussion

The benefits of the ADPREP algorithm compared to other EP detection methods are multifold. Firstly, the algorithm does not assume a particular EP waveform, which means it does not discriminate towards or against any particular shapes – any neural response that is common to both stimulation polarities can be labeled as an EP. Secondly, since the algorithm is simply based on correlation, it is potentially applicable to any kind of bipolar stimulation that allows for polarity reversal, when the recording electrodes are at a close enough proximity to the stimulation electrodes to register the polarity reversal of the stimulus artifact. I.e., the ADPREP algorithm is not restricted to the deep brain stimulation during deep brain recordings that was exemplified in this paper. Thirdly, the user-defined constants and thresholds allow the user to tailor the algorithm to their particular application, e.g., by selecting the moving correlation window size(s) to match the frequency and duration of the expected EPs, or lowering or raising the amplitude threshold to reduce the false negative or false positive rates, respectively. Finally, the automated algorithm vastly improves efficacy and minimizes human bias by reducing the influence of the user, which sets each study up for more reliable further analysis.

As with any method, the ADPREP algorithm also comes with limitations. Firstly, because the cathode acts as the active electrode in the stimulating pair, and the anode as the returning electrode, the polarity reversal technique effectively leads to a slight shift in stimulation location, corresponding to the distance between the stimulated electrode pairs (5 mm, in our case of using the macro-contacts of the Ad-Tech MM16C sEEG leads). Therefore, the final averaged response of the two stimulation types is, in fact, an average of stimulations at two slightly different locations, which may lead to timing or amplitude differences between the two recordings.

Furthermore, in its current version, the algorithm is designed to detect one EP per recording and stimulation, and in the case of multiple possible EPs, the one with the largest P2P amplitude is selected. These assumptions could be addressed in future versions, e.g., by redefining how the algorithm should select between multiple possible EPs and/or allowing for the labeling of more than one EP per recording. Moreover, while the method should work for any set of stimulating electrodes that allow for polarity reversal and with recording electrodes that are proximal enough to experience a reversed polarity of the stimulus artifact, it has so far only been tested in deep brain sEEG recordings during DBS. Tests with other stimulating and recording modalities, such as EEG, must be conducted before it can be conclusively stated which applications the ADPREP algorithm is compatible with.

## Conclusion

Our algorithm for automated detection of polarity reversed evoked potentials (ADPREP) utilizes polarity reversal of the stimulating electrode pair to distinguish neural responses from stimulus artifacts and other noise. It has proven useful in detecting DBS EPs in deep brain nuclei (basal ganglia and thalamus) and has promising applications in other stimulation and recording modalities that allow for polarity reversal of the recorded stimulus artifact. While the algorithm can be further developed, e.g., to allow for detection of multiple EPs per recording, it has already proven successful in labeling EPs of varying shapes and sizes, as early as a fraction of a millisecond after stimulation, in a range of stimulus frequencies and stimulation-recording pairs in deep brain nuclei, in more than two dozen pediatric patients with dystonia – even under large stimulus decay artifacts. As such, it is a valid option to improve efficacy and minimize human bias, setting up for more reliable analysis and conclusions from the datasets.

## Data Availability

All data in the present work are confidential.

## Acknowledgements

We thank our subjects and their parents for participating in this study, and are appreciative of the significant efforts of the inpatient nursing and surgical staff at both CHOC and CHLA. We also thank Jennifer MacLean, Allison Przekop, Teresa Serna-Fonseca, and Diana Ferman for their assistance with neurologic examinations and electrical stimulation, Estefanía Hernandez-Martín, Jaya Nataraj, Maral Kasiri, Alireza Mousavi, Sina Javadzadeh, Enrique Argüelles and Ruta Deshpande for helping with subject testing and data collection, and Rachel Davis for manuscript editing.

